# Gene expression and alternative splicing analysis in a large-scale Multiple Sclerosis study

**DOI:** 10.1101/2024.08.16.24312099

**Authors:** Müge Sak, Julia H. Chariker, Juw Won Park, Eric C. Rouchka

## Abstract

**Background:** Multiple Sclerosis (MS) is an autoimmune neurodegenerative disease affecting approximately 3 million people globally. Despite rigorous research on MS, aspects of its development and progression remain unclear. Understanding molecular mechanisms underlying MS is crucial to providing insights into disease pathways, identifying potential biomarkers for early diagnosis, and revealing novel therapeutic targets for improved patient outcomes.

**Methods:** We utilized publicly available RNA-seq data (GSE138614) from post-mortem white matter tissues of five donors without any neurological disorder and ten MS patient donors. This data was interrogated for differential gene expression, alternative splicing and single nucleotide variants as well as for functional enrichments in the resulting datasets.

**Results:** A comparison of non-MS white matter (WM) to MS samples yielded differentially expressed genes involved in adaptive immune response, cell communication, and developmental processes. Genes with expression changes positively correlated with tissue inflammation were enriched in the immune system and receptor interaction pathways. Negatively correlated genes were enriched in neurogenesis, nervous system development, and metabolic pathways. Alternatively spliced transcripts between WM and MS lesions included genes that play roles in neurogenesis, myelination, and oligodendrocyte differentiation, such as brain enriched myelin associated protein (*BCAS1*), discs large MAGUK scaffold protein 1 (*DLG1*), KH domain containing RNA binding (*QKI*), and myelin basic protein (*MBP*). Our approach to comparing normal appearing WM (NAWM) and active lesion (AL) from one donor and NAWM and chronic active (CA) tissues from two donors, showed that different IgH and IgK gene subfamilies were differentially expressed. We also identified pathways involved in white matter injury repair and remyelination in these tissues. Differentially spliced genes between these lesions were involved in axon and dendrite structure stability. We also identified exon skipping events and spontaneous single nucleotide polymorphisms in membrane associated ring-CH-type finger 1 (*MARCHF1*), UDP glycosyltransferase 8 (*UGT8*), and other genes important in autoimmunity and neurodegeneration.

**Conclusion:** Overall, we identified unique genes, pathways, and novel splicing events affecting disease progression that can be further investigated as potential novel drug targets for MS treatment.

## INTRODUCTION

Multiple Sclerosis (MS) is an autoimmune neurodegenerative disease of the central nervous system (CNS) that affects about 3 million people worldwide [1]. Magnetic resonance imaging (MRI) of the brain and spinal cord is used to identify characteristic lesions, while evidence of intrathecally-produced immunoglobulin G (IgG) via lumbar puncture is used as a diagnostic test for MS [2]. Vitamin D deficiency and Epstein-Barr virus (EBV) infections have been consistently linked to MS diagnosis [3, 4]. Although MS is not an inherited disease, genetic susceptibility to MS is known [5, 6]. This includes 233 risk loci identified in genome-wide association studies [7]. Thirty of those loci are on the major histocompatibility complex (MHC) suggesting a primary role in autoimmunity [8].

MS is a complex disease with an unclear and multifactorial origin and lesion pathology. Lesions are characterized by inflammation, demyelination, and neurodegeneration of the CNS. These lesions differ in distribution and composition both among different patients and within individuals [9]. Their specific locations often correlate with clinical symptoms, leading to significant clinical variability [10, 11]. Several biological processes take place during the progression of MS: the autoreactive T-cell and B-cell aggregation in the CNS, secretion of cytokines, damage to myelin sheath, and oligodendrocytes [12-16]. The timing of these processes resulting in neurodegeneration is important for therapeutic strategies [17]. Various imaging strategies have tested variables relevant to timing the disease processes, leading to staging systems for MS [10, 14, 18]. However, there is no standard method for the classification of the lesions. The histological staging system for the post-mortem tissues includes inflammatory cells, glial cells, axonal loss, and myelin staining [18, 19].

The damage to myelin sheaths and oligodendrocytes occurs as a result of inflammation, disruption of the blood-brain barrier (BBB), and immune cell infiltration to CNS [20]. Axons are relatively preserved in the early stages of the disease; however, as the disease progresses, irreversible axonal damage develops [21]. Early studies of MS research focused on the intrathecal immunoglobulin synthesis [22-24]. With the discovery of the risk alleles in the MHC region and the novel findings of T-cells, MS is thought to be T-cell-driven [25, 26]. However, with the recent discovery of the efficacy of B-cell depletion therapies and better technologies to study the adaptive immune system, interest in antibodies in MS has arisen [27] and the profound involvement of the adaptive immune system and B-cells in disease pathology has been shown [28].

Here we utilized publicly available RNA-seq data from postmortem white matter (WM) tissues from 5 non-MS and 10 MS patient donors [29]. The previous analysis with this dataset identified lesion-specific molecular signatures, protein complex networks, and transforming growth factor beta receptor 2 (*TGFβ-R2*) as a central hub [30]. Since each lesion had a high complexity of molecular pathways, the simple molecular categorization of lesion types was not feasible, indicating a dynamic process of lesion evolution [31]. In order to extract additional information from this dataset, we employed differential expression, differential splicing, and variant analysis both among and within donors. This approach identified genes that may be involved in the progression of inflammation. With the non-MS vs MS tissue comparisons, we identified over 6,000 differentially expressed genes (FDR < 0.05). The most differentially expressed genes were involved in inflammation, immune system processes, and cell signaling. In comparing WM to different lesion types, we investigated genes that follow similar expression patterns as the inflammation level in the tissues. The genes that show a positive correlation with inflammation were enriched in immune system processes, immune response and response to stimulus and enriched in KEGG pathways such as cytokine-receptor interaction. The negatively correlated genes were enriched in neurogenesis and nervous system development in biological processes and pathways. Donor-specific differential expression analysis showed that immunoglobulin gene subfamilies were differentially expressed. Finally, differential splicing and variant analyses identified genes that play a role in dysregulation in immune system, metabolism, and axon/dendrite structure stability. Currently, there is no cure for MS. Disease-modifying therapies help recovery from attacks, modify the course of the disease, and manage symptoms. Our comprehensive analysis of differential expression and splicing of genes in MS patients demonstrates the role of previously recognized genes, as well as novel pathways. These results can lead the way to identifying novel genes and drug targets for reversing the damage, preventing MS progression, and helping patients with disease management.

## METHODS

### Data access and characteristics

The publicly available RNA-seq dataset was retrieved from the GEO repository (accession GSE138614) [32]. The original study that generated the data was approved by the local ethics committee of the original authors. The lesion classification method was explained in El Kajaer et. al [30]. Namely, normal-appearing white matter (NAWM), active (AL), chronic active (CA), inactive (IL), and remyelinating (RL) lesions were characterized by myelin integrity and the inflammatory state [18]. For the comparisons of NAWM tissues with AL or CA lesions within the same donors, the accession numbers are shown in Table 1.

**Table 1:** Sequence Read Archive accession numbers of the sample-specific comparisons. AL: active lesion; CA: chronic active lesion NAWM: normal-appearing white matter.

| Sample ID | Lesion Type | SRA Accession Number |
| --- | --- | --- |
| S6 | AL | SRR10248474 |
| S6 | AL | SRR10248477 |
| S6 | AL | SRR10248479 |
| S6 | NAWM | SRR10248426 |
| S6 | NAWM | SRR10248446 |
| S6 | NAWM | SRR10248448 |
| S14 | CA | SRR10248408 |
| S14 | CA | SRR10248409 |
| S14 | CA | SRR10248412 |
| S14 | CA | SRR10248440 |
| S14 | CA | SRR10248443 |
| S14 | NAWM | SRR10248423 |
| S14 | NAWM | SRR10248433 |
| S14 | NAWM | SRR10248449 |
| S14 | NAWM | SRR10248452 |
| S9 | CA | SRR10248415 |
| S9 | CA | SRR10248420 |
| S9 | CA | SRR10248439 |
| S9 | NAWM | SRR10248414 |
| S9 | NAWM | SRR10248430 |
| S9 | NAWM | SRR10248434 |
| S9 | NAWM | SRR10248444 |
| S9 | NAWM | SRR10248447 |

### Differential expression analysis

Reads from 73 MS samples from 10 MS donors and 25 samples from 5 non-MS donors were mapped to the human genome (hg38) using the Spliced Transcripts Alignment to a Reference (STAR) aligner (version 2.6) [33]. Raw gene counts were determined using HTSeq-count (version 0.10.0) [34]. Raw counts were then normalized using the relative log expression method and filtered to exclude genes with fewer than 10 counts across all samples. Differential expression analysis was performed with DESeq2 [35] using a negative binomial regression model to analyze pairwise comparisons. Statistical significance was determined based on a false discovery rate (FDR) cutoff of 0.05.

### Inflammation level analysis

STEM is an algorithmic approach for clustering, comparing, and visualizing short-time series gene expression [36]. STEM selects a set of distinct and representative temporal expression profiles that are independent of the data. Each gene is assigned to the model profile that most closely matches the expression profile, determined by the correlation coefficient. A permutation test determines the assignments of genes to model profiles. The significance of each model profile is calculated by the number of assigned genes under the true ordering of time points compared to the average number assigned to the model profile [36]. STEM was utilized to investigate the correlation between gene expression and tissue inflammation. Instead of using time series samples, the inflammation level of each sample was used to determine the genes that showed positive and negative correlations.

### Alternative splicing analysis

Replicate multivariate analysis of transcript splicing (rMATS v3.2.5) [37] was used to identify differentially spliced genes. rMATS employs a modified generalized linear mixed model to identify splicing from RNA-seq data with replicates. Using both splice junction and exon body read counts as input, rMATS computes the *percent spliced in* (PSI) and FDR for five different major types of splicing events: skipped exons (SE), mutually exclusive exons (MXE), retained introns (RI), and 5′ and 3′ alternative splice sites (A5SS and A3SS). Motif enrichment was analyzed in the proximity of alternatively spliced exons using rMAPS2 [38, 39]. rMAPS2 analyzes differential alternative splicing data obtained from rMATS and then graphically displays enriched motif sites.

### Functional enrichment analysis

Gene ontology biological processes (GO:BP) enrichments were determined via hypergeometric testing using clusterProfiler [40] for differentially expressed and differentially spliced genes. Gene set enrichment analysis (GSEA 4.3.2) was utilized on pre-ranked differentially expressed genes within sample lesions comparisons [41].

### RNA-seq short variant discovery

RNA-seq short variants were identified using GATK’s workflow as a guide [42]. Alignment files were preprocessed using Picard’s (v_2.25.6) AddOrReplaceGroups and GATK’s (v-4.2.0.0) tool MarkDuplicates. SplitNCigarReads formatted the RNA-seq alignments for use in HaplotypeCaller. Base quality and recalibration were performed to correct for systematic errors in base quality scores prior to using HaplotypeCaller for variant calls. SNVs were filtered based on Fisher Strand values (FS > 30.0) and Qual by Depth values (QD < 2.0).

## RESULTS

### Differentially expressed genes in non-MS versus MS samples

We identified 6,461 differentially expressed genes between MS and non-MS tissue samples (FDR < 0.05) (Figure 1). Among these, the joining chain of multimeric IgA and IgM (*JCHAIN/IGJ*) was identified as the most significant differentially expressed gene in white matter tissue samples from MS donors. This finding aligns with Christensen et al.’s study [43], which reported increased *JCHAIN* expression in cerebrospinal fluid samples from MS patients but not in blood samples, suggesting a specific role in CNS inflammation. Other genes with high expression in MS patient samples were pro-inflammatory, such as immunoglobulin lambda like polypeptide 5 (*IGLL5*), involved in B-cell development [44], collagen type VIII alpha 1 chain (*COL8A1*) [45] and signaling lymphocytic activation molecule family member 7 (*SLAM7*), which regulates B cells and adaptive immunity and affects susceptibility to CNS autoimmunity [46]. We also identified upregulation of paralemmin 3 (*PALM3*) and hepatocyte growth factor (*HGF*) genes that are involved in response to inflammation [47]. Additionally, overexpressed IKAROS family zinc finger 3 (*IKZF3*), MER proto-oncogene, tyrosine kinase (*MERTK*), and Fc receptor-like 5 (*FCRL5*), all of which have risk alleles associated with MS and other autoimmune diseases [48-52] (Figure 1A). The most downregulated genes were primarily involved in various signal transduction and biosynthesis pathways, including tyrosine 3-monooxygenase/tryptophan 5-monooxygenase activation protein theta (*YWHAQ*), protein tyrosine phosphatase domain containing 1 (*PTPDC1*), RAB guanine nucleotide exchange factor 1 (*RABGEF1*), 1-acylglycerol-3-phosphate O-acyltransferase 4 (*AGPAT4*), and SH3 domain binding protein 5 (*SH3BP5*) (Figure 1A).

**Figure 1:**
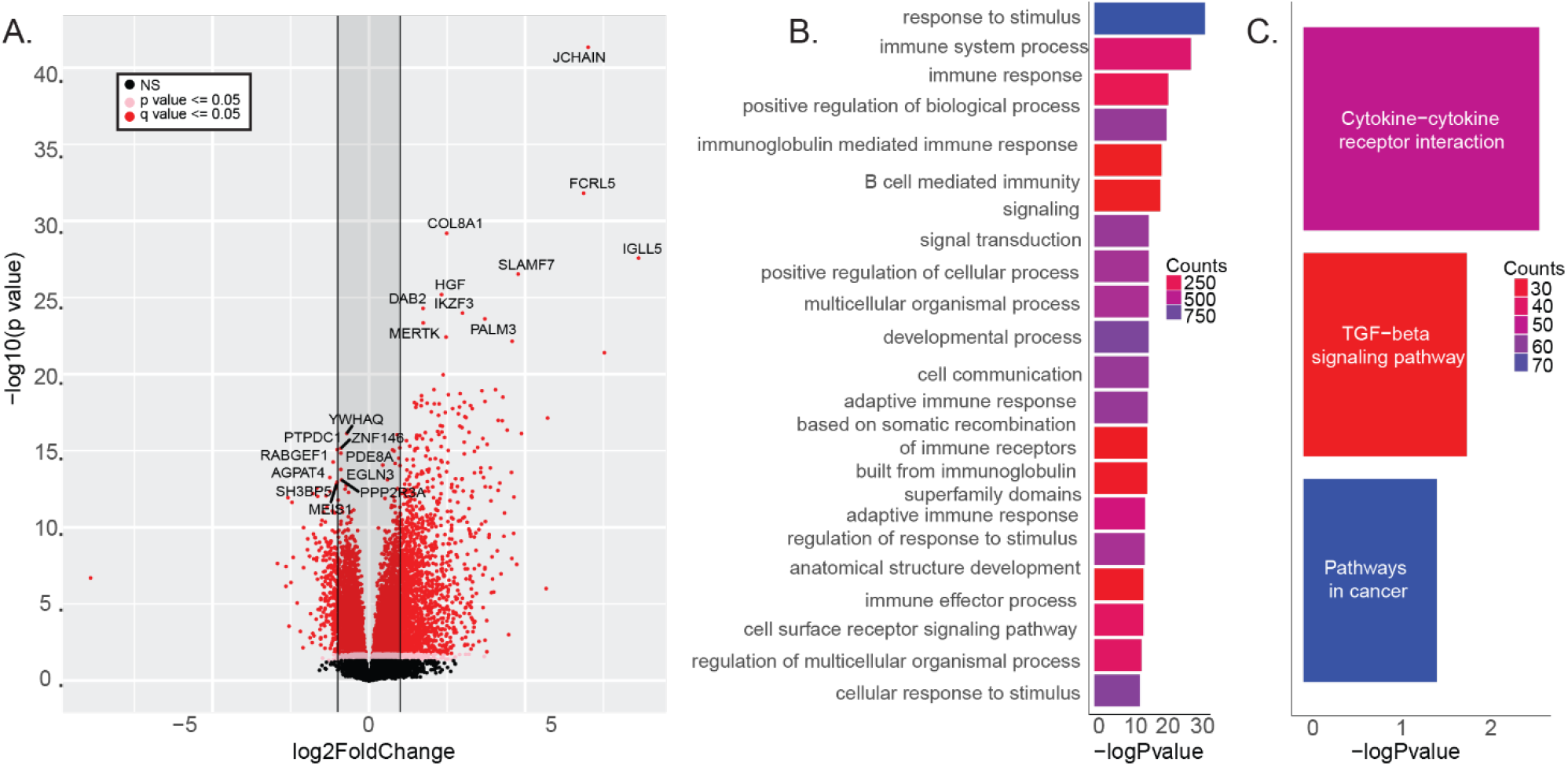
Differential expression of WM tissues from MS and non-MS donors. **A:** Volcano plot showing differentially expressed genes between MS and non-MS donor tissue samples. **B:** Enriched gene ontology biological processes and **C:** KEGG pathways from the differentially expressed genes.

Functional enrichment analysis revealed that these differentially expressed genes were significantly involved in stimulus-response, immune system, and cell signaling functions (Figure 1B), as well as proinflammatory cytokine-cytokine receptor interaction and TGF-beta signaling pathways (Figure 1C).

### Investigation of the genes correlating with the tissue inflammation level

We identified 2,886 genes that showed a positive correlation with the inflammation levels in the tissue (Figure 2A) and 647 genes showed a negative correlation (Figure 2B). Positively correlated genes enriched in biological pathways were similar to the differential expression analysis, such as immune system process, immune response, and response to stimulus (Figure 2C). Additionally, we identified viral protein interaction with cytokine receptors, vitamin digestion and absorption, and natural killer cell-mediated cytotoxicity pathways associated with inflammation levels in MS tissues. Genes showing expression levels that are negatively correlated with the tissue inflammation were enriched in the cellular anatomical entity, morphogenesis, and neurogenesis biological processes and metabolic pathways (Figure 2D).

**Figure 2:**
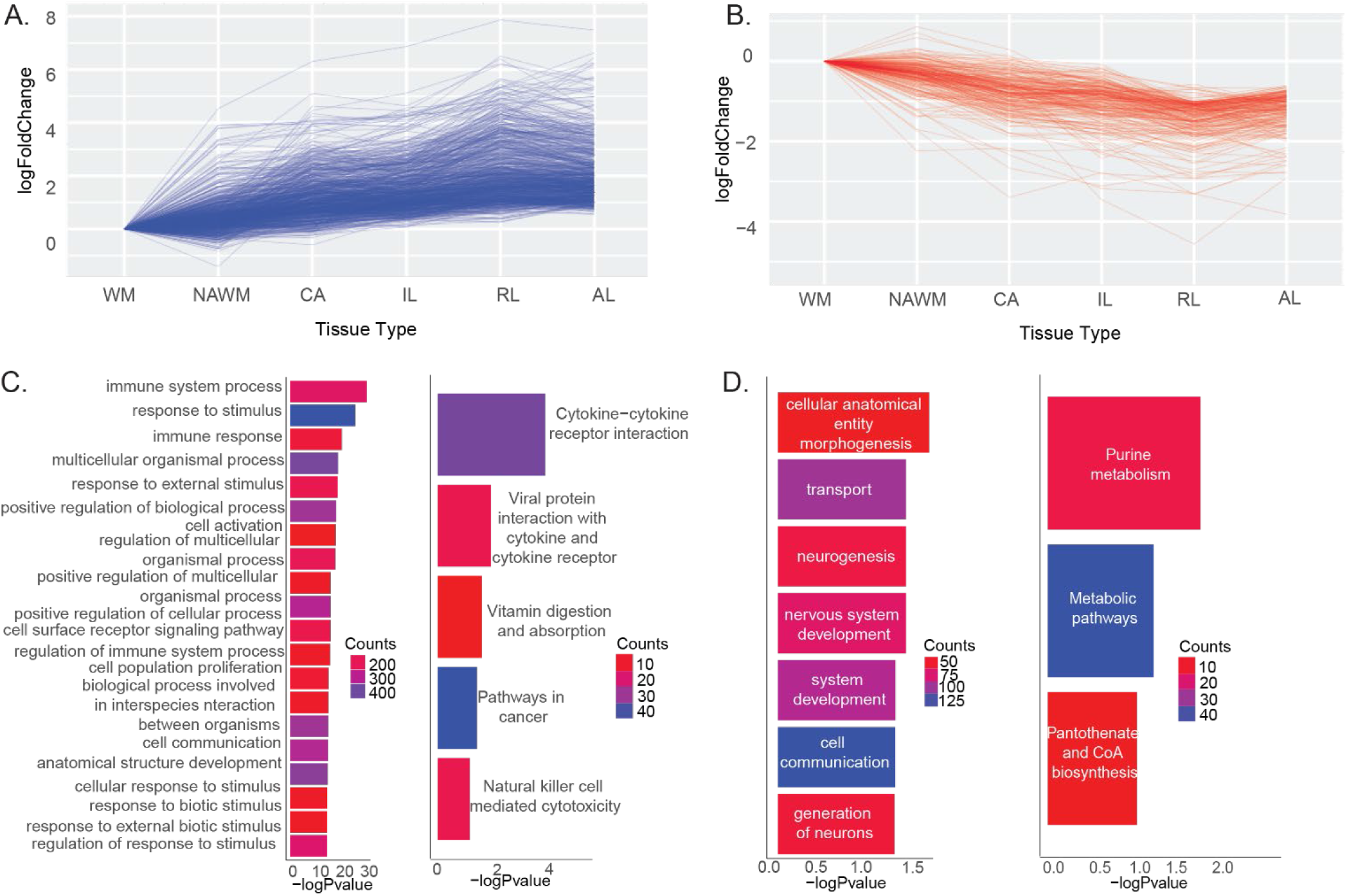
Genes showing A. positive and B. negative correlation in expression changes with tissue inflammation levels. **C:** Enriched gene ontology biological and KEGG pathways for genes showing positively correlated expression levels with inflammation. **D:** Enriched gene ontology biological and KEGG pathways for genes showing negatively correlated expression levels with inflammation.

### Differential splicing and differential expression of RNA binding proteins (RBPs)

We identified differentially spliced genes in each tissue compared to non-MS WM with a total of 2,721 genes alternatively spliced in MS tissues. The intersections of alternative splicing events across comparisons are illustrated in Figure 3A. The representative motif bindings for each type of splicing (Supplemental Figure 1) and differentially expressed RNA Binding Proteins (RBPs) that bind to these motifs in MS lesions are shown in Figure 3B. These suggest the mechanism of alternative splicing may result from differentially expressed of RBPs.

**Figure 3:**
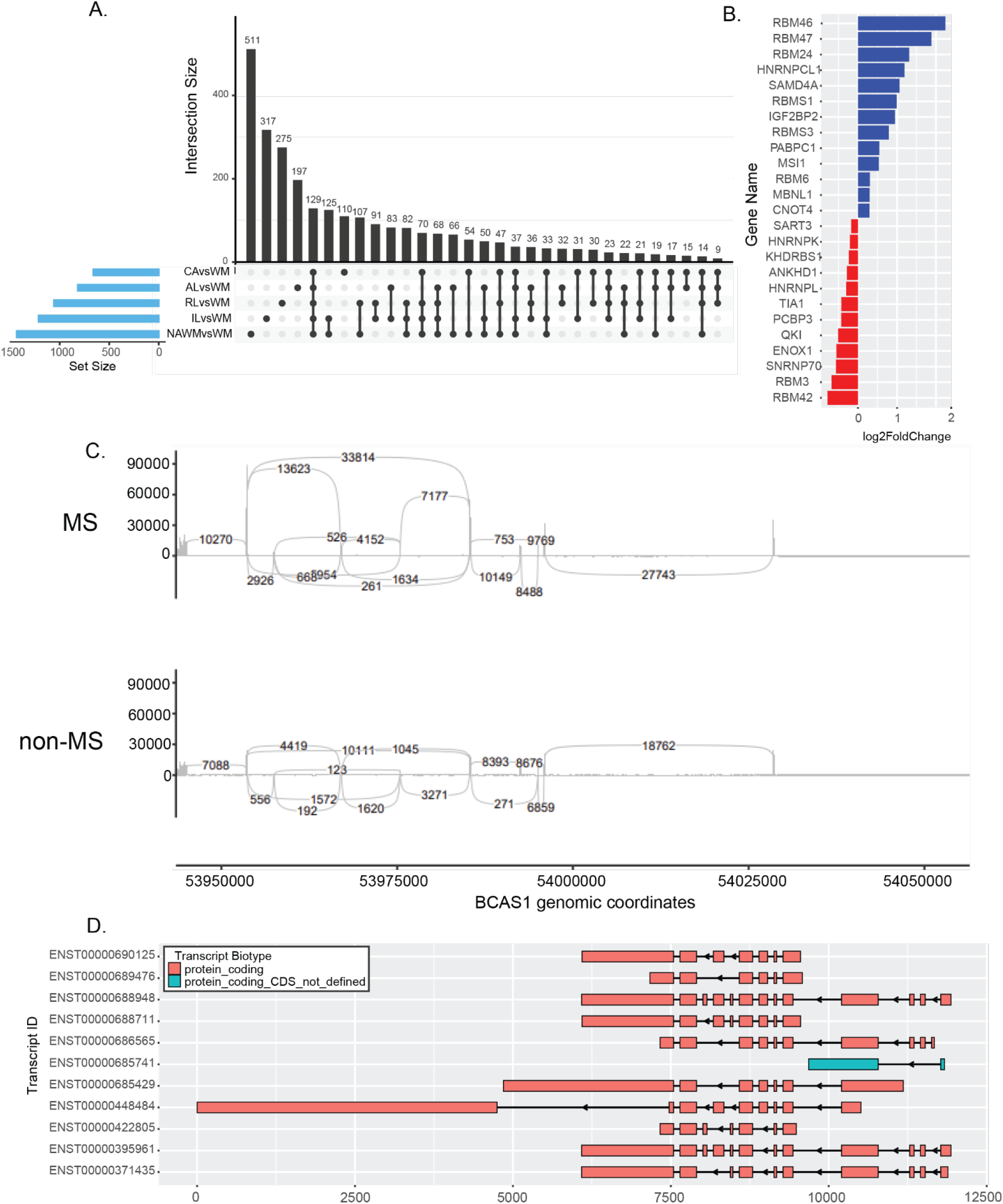
Differentially spliced genes in all tissues from MS donor samples compared to non-MS samples. **A:** Upset graph showing the number of alternatively spliced genes in each category of tissues and intersections. **B:** Differentially expressed RNA binding proteins that are involved in alternative splicing. **C:** Sashimi plot of *BCAS1* gene that is differentially spliced in MS and non-MS tissues. **D:** Known transcripts of *BCAS1* gene.

Key genes with alternative splicing included myelin basic protein (*MBP*), DEAD-box helicase 5 (*DDX5*), KH domain containing RNA binding (*QKI*), and discs large MAGUK scaffold protein 1 (*DLG1*) (Table 2).

**Table 2:**
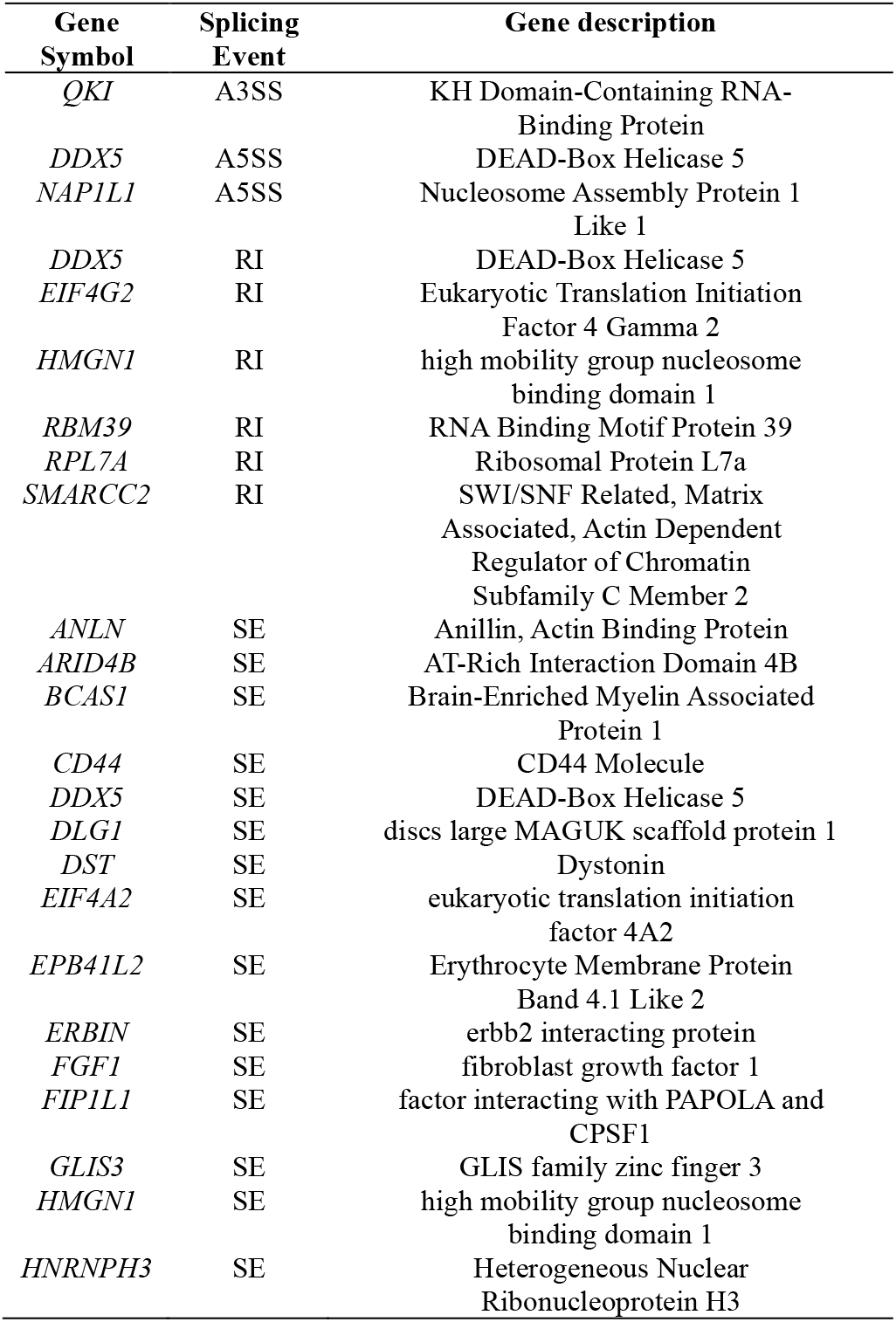
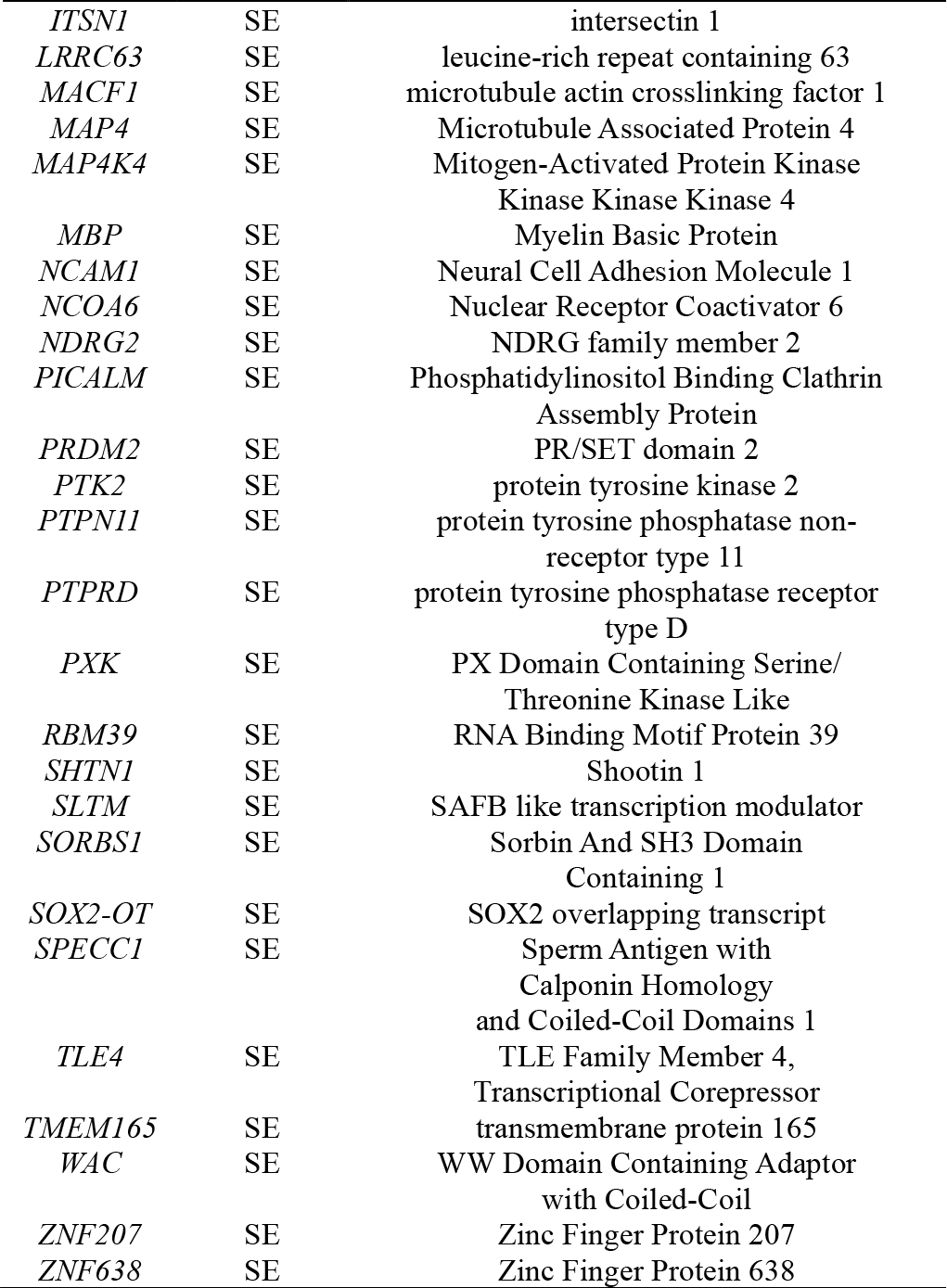
Differentially spliced genes in the MS patient samples. A3SS: alternative 3’ splice site; A5SS: alternative 5’ splice site: RI: retained intron; SE: skipped exon.

Recent research indicates that deletion of *QKI* isoforms in oligodendrocytes leads to severe CNS hypomyelination accompanied by tremors [53]. Furthermore, *DDX5*, which exhibited A5SS, RI, and SE in MS tissues, influences *MBP* expression [54]. *DLG1* plays a crucial role in lymphocyte activation [55] and has splice variants that regulate p38-dependent and independent effector functions in CD8+ T cells [56]. We also found MS patients had significant SE events in brain enriched myelin associated protein (*BCAS1*) that can result in expression of alternative isoforms (Figure 3C-D). BCAS1 is important for early myelination [57] and carries a known MS risk allele rs2585447 [7].

### Comparison of NAWM with AL or CA within donors

We examined differential expression and splicing within the same donors to minimize inter-donor variation and closely analyze the CA and AL lesions by comparing them to the NAWM from the same donor. This approach enabled a detailed and specific examination of the molecular differences in the lesion areas of patient brains. A comparison of NAWM and AL showed increased expression of *IGKV4-1, IGHV1-2*, and *IGHV4-59* as well as local inflammation marker *CD163* and interleukin 5 receptor alpha (*IL5RA*) (Figure 4A, Supplemental Table 1). *CD24*, which carries polymorphisms linked to the progression of autoimmune disorders [58] and *JCHAIN* genes were differentially expressed in both AL and CA comparisons to NAWM for three donors. The *IGHV3-30* gene was overexpressed in donor S9 for the CA and NAWM comparison. (Figure 4B, Supplemental Table 2). For donor S14, we found the immunoglobulin genes *IGKC, IGHV3-7, IGHG1, IGHV4-39*, and *IGHG3* were overexpressed in CA lesions compared to NAWM (Figure 4C. Supplemental Table 3). In addition to *CD24*, the CA to NAWM comparison in the S9 and S14 donors showed overexpression of hepatocyte growth factor (*HGF*), extracellular matrix protein (*ECM2*), complement 6 (*C6*), ceruloplasmin (*CP*), and insulin growth factor binding protein 7 anti-sense RNA 1 (*IGFBP7-AS1*) (Figure 4B-C, Supplemental Tables 2-3). This suggests these genes may have specific roles in CA lesions.

**Table 3:**
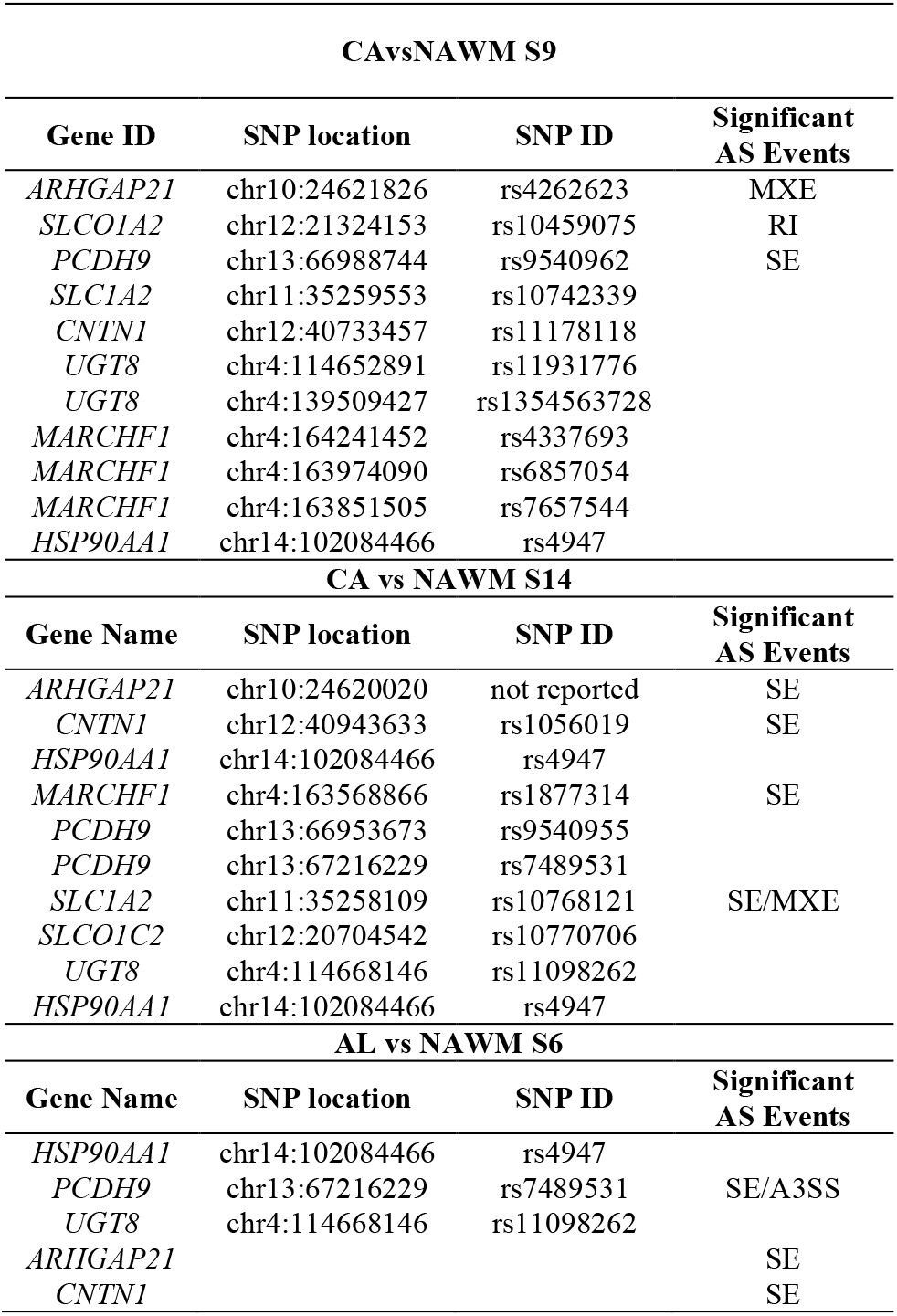
Common SNPs and alternative splicing events in AL and CA tissue comparisons to NAWM. A3SS: alternative 3’ splice site; MXE: mutually exclusive exon; RI: retained intron; SE: skipped exon.

**Figure 4:**
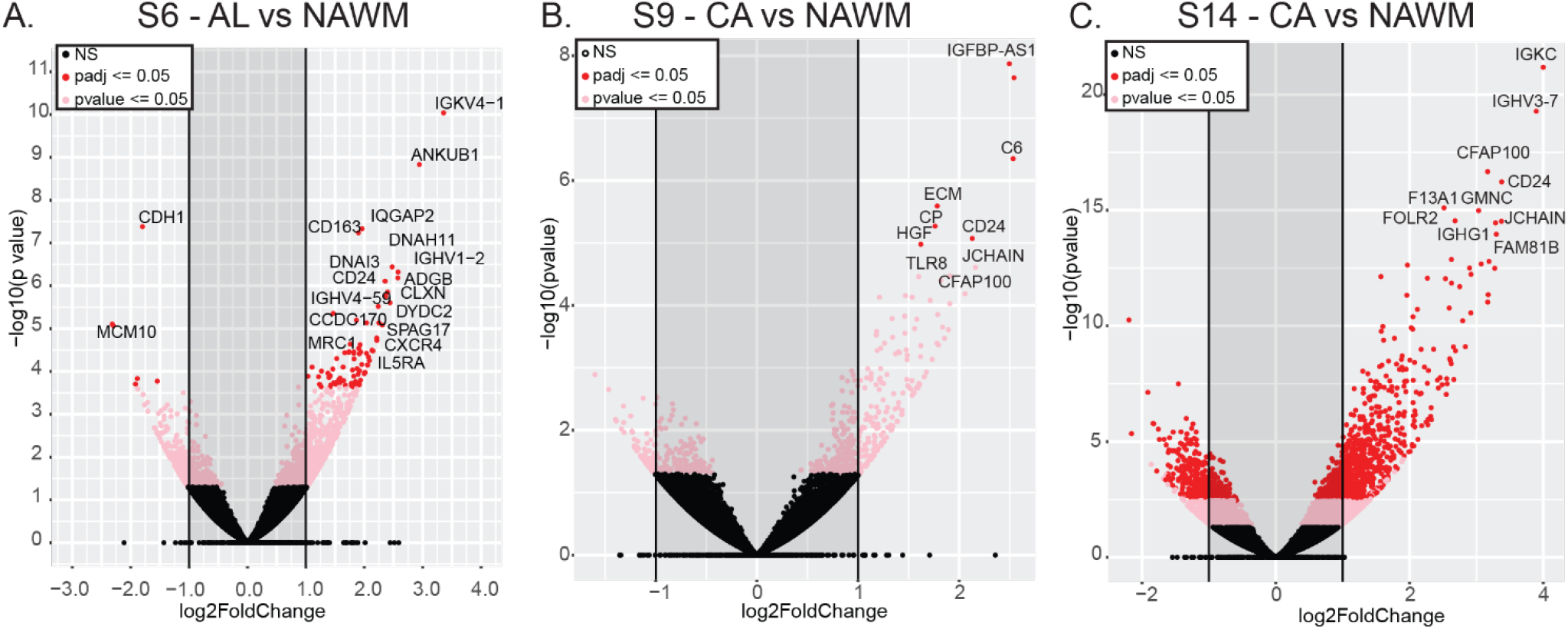
Differentially expressed genes in different tissue types from the same donors. Volcano plots showing differentially expressed genes in **A:** AL compared to NAWM in donor S6, CA compared to NAWM in **B:** donor S9 and **C:** S14.

Differentially expressed AL genes were enriched in cilium movement, mainly through the upregulation of cilia and flagella associated protein 100 (*CFAP100*), cilia and flagella associated protein 45 (*CFA45*), and dynein axonemal heavy chain 5 (*DNAH5*) genes. Negative enrichment of postsynaptic membrane potential regulation indicates a substantial damage to neurons in which synaptic failure eventually leads to brain network alterations and contributes to disabling MS symptoms and disease progression [59] (Supplemental Table 4). Splice variants in AL compared to NAWM lesions were mainly enriched in metabolic processes. We found significant disruption of cellular localizations by alternatively spliced genes (Supplemental Figure 2A) that could be involved in disrupted interleukin receptor localizations in MS tissues [60].

Gene set enrichment analysis for differentially expressed genes in CA and NAWM showed B cell-mediated immunity and axoneme assembly were the most positively enriched pathways. Negatively enriched short-chain fatty acid and acetyl-COA metabolic processes have been shown to have an immunomodulatory potential in MS [61] (Supplemental Tables 5-6). Differentially spliced genes in the CA lesions compared to the NAWM from the same patients were mostly enriched in cellular component organization, nervous system development, and cell morphogenesis pathways, indicating possible pathogenicity by alternative splicing contribution to mitochondrial and metabolic dysfunction of the CA lesions (Supplemental Figure 2B).

### Spontaneous single nucleotide variations and alternative splicing

A comparison of the single nucleotide variations (SNVs) in MS and non-MS samples did not match any known SNV markers of MS. This is likely due to the small number of donors. We then compared SNVs in AL and CA lesions to NAWM lesions in the S6, S9 and S14 donors and identified a common synonymous variation (rs4947) in the heat shock protein 90 alpha family class A member 1 (*HSP90AA1*) gene across all comparisons. We also identified SNVs on rho GTPase activating protein 21 (*ARHGAP21*), contactin 1 (*CNTN1*), solute carrier family 1 member 2 (*SLC1A2*), UDP glycosyltransferase 8 (*UGT8*), solute carrier organic anion transporter family member 1C1 (*SLCO1C1*), protocadherin 9 (*PCDH9*), and membrane associated ring-CH-type finger 1 (*MARCHF1*). Between CA and NAWM tissues, *ARHGAP21* had significant SE events in both the S9 and S14 donors, as well as in the AL lesions of the donor S6. *SLC1A2* had significant SE events in S14. *CNTN1* also showed an SE variant in S14 CA lesions and S6 AL lesions. The same events for *SLC1A2* and *CNTN1* were found in S9 but were not determined to be significant. *PCDH9* carried spontaneous SNVs in all comparisons and had a significant SE event in both AL and CA lesions from S6 and S9 donors compared to their NAWMs. (Table 3).

MARCHF1 is an E3 ubiquitin ligase and MARCH1-mediated ubiquitination of MHC II impacts the MHC I antigen presentation pathway [62]. *MARCHF1* carries a known MS risk variant rs72989863. We identified different spontaneous SNVs that are upstream variants in CA lesions from both S9 and S14 donors. The S14 donor also had a significant SE variant in CA lesions (Figure 5A). The same region was also skipped in CA lesions from S9 but were not significant. In donor S6, the comparison of AL and NAWM tissues yields the same SE event on *MARCHF1*. This region corresponds to the exon 4 of the ENST00000514618 transcript and this SE event may result in the expression of the ENST0000503008 transcript (Figure 5B). These two transcripts translate into two different MARCHF1 protein isoforms that may alter its E3 ubiquitin ligase activity (uniport IDs Q8TCQ1 and D6RGC4) [63, 64]. However, it is essential to note that our analysis utilized short-read sequencing data which does not allow identification of specific transcripts.

**Figure 5:**
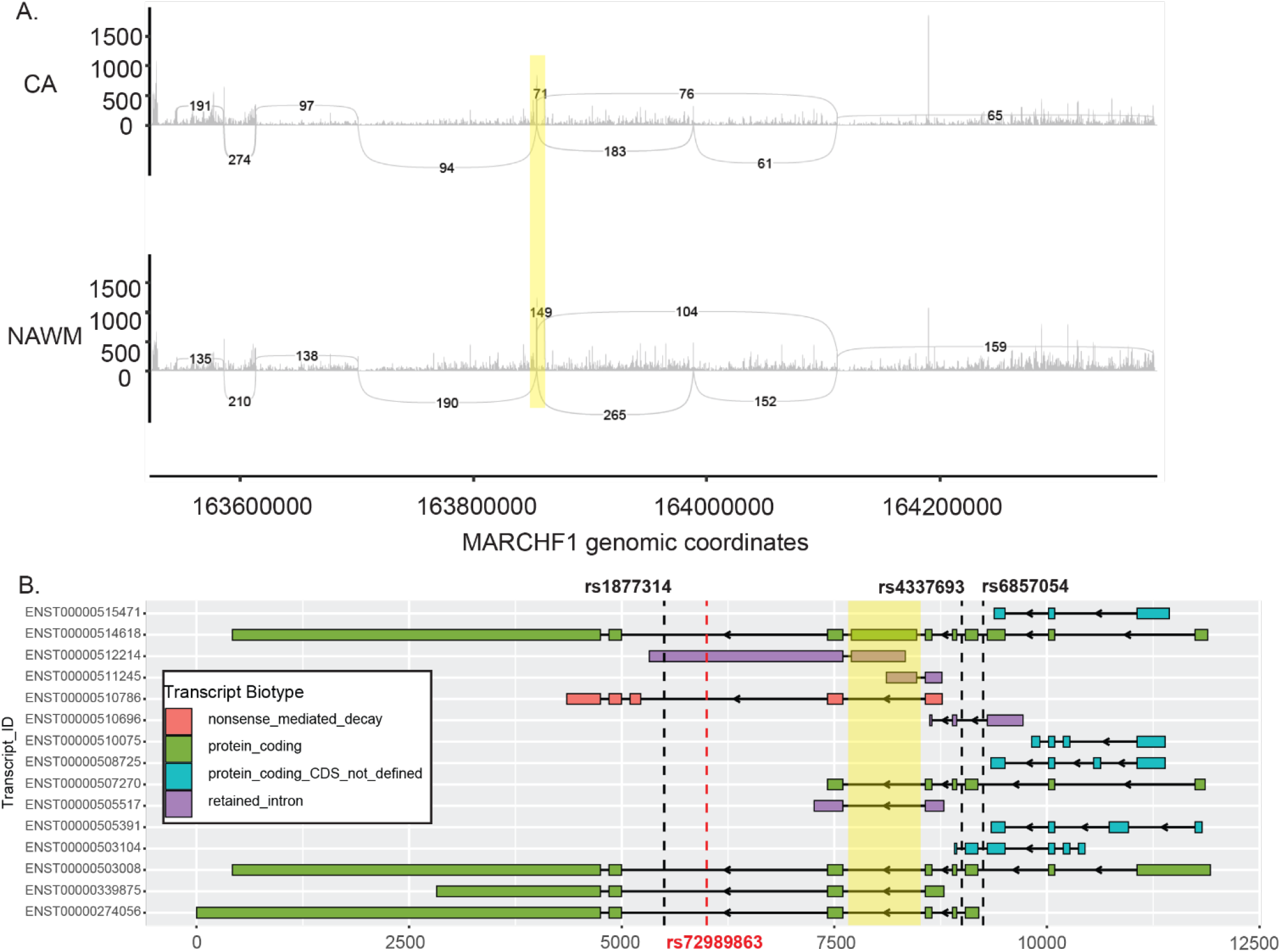
Sashimi plot of the *MARCHF1* gene that is differentially spliced in CA lesions. Skipped exon 4 is highlighted in yellow. **B:** Known transcripts of MARCHF1 gene. Known MS risk allele is shown in red (rs72989863) and the identified spontaneous SNPs are shown in black. Skipped exon 4 is highlighted in yellow.

## DISCUSSION

MS is highly complex and variable, both among patients and the stages of the disease [65]. Understanding the molecular processes through disease progression and identifying molecular signatures of the specific pathology of the MS lesions is crucial for understanding the disease development and progression, as well as for patient stratification for prognosis, predicting treatment response, and identifying treatment strategies [66]. The dataset we utilized has a large number of samples from different types of MS lesions which allowed us to investigate differences in these lesions at the level of differential gene expression, alternative splicing, and SNVs. Identifying gene expression patterns correlating with tissue inflammation can be used as a marker for lesion stage determination. However, with a large number of samples, greater variability makes it difficult to cluster samples using their expression profiles (e.g. PCA). The source of this variability may be the result of sample collection from different parts of the brain [67].

Alternative splicing is one of the main mechanisms affecting the expression of genes and gene isotypes. A recent study showed that there are a large number of alternative splicing variants in MS that are not linked to the differential expression [68] and there are splice variants found to be associated with known risk alleles [69]. Many of the alternatively spliced genes we identified play roles in MS-related pathways. Deletion of *QKI* isoforms in oligodendrocytes leads to severe CNS hypomyelination accompanied by tremors [53]. *DLG1* plays a crucial role in lymphocyte activation and has identified spliced variants that regulate p38-dependent and independent effector functions in CD8+ T cells [56]. *BCAS1* is highly expressed in oligodendrocytes and plays role in demyelination in MS [57].

We discovered *BCAS1*, which carries a known MS risk allele rs2585447 [7], had a splice variant in MS patient samples. Our analysis also identified differential splicing of *MBP* and *MOBP* in different types of MS lesions that show variable expression levels in MS [70, 71]. Studying these alternative isoforms more thoroughly could enhance our comprehension of how MS progresses and how we might be able to halt or reverse it.

Although we had limited samples for the tissue type comparisons within donors, our approach in investigating the differential expression and differential splicing of CA vs NAWM and AL vs NAWM lesions resulted in the identification of specific pathways such as fatty acid and acetyl COA metabolic processes, as well as cellular components and cytoskeleton organization. Lipids are not solely involved in the formation of the myelin sheath but are found to be important components of cell signaling, communication, and transport in CNS and have been shown to be relevant players in neuroinflammation and neurodegeneration [72]. We identified specific pathways that are affected in specific stages of MS lesions, which can be markers for the staging of these lesions, as well as the selection of treatment strategies. They are also the basis for further research in reversing damage and treating disability symptoms of MS. The inflammatory activities in lesions are shown to result in disease progression without any new lesion formation [73]. In AL, we identified markers for neuronal damage by changes in cilia and postsynaptic membrane potential pathways. Targeting these specific proteins and pathways, may block the silent progression and prevent further disability development in the patients.

Comparison of NAWM tissues with CA or AL within patients identified spontaneous SNVs that may be associated with alternative splicing in specific MS lesions. Among the genes we identified, ARHGAP21 is involved in the signaling for synaptic homeostasis and axon/dendritic transport regulation [74]. Serum CNTN1 concentrations are associated with the MS progression in RRMS patients [75]. Solute carrier family 1 member *SLC1A2* is expressed in astrocytes, neurons, and axonal terminals and upregulated in MS. It regulates glutamate concentrations in the CNS preventing excitotoxicity [76, 77]. UGT8 plays an important role in remyelination by mediating the major myelin lipid galactosylceramide [78]. MARCHF1 is an E3 ubiquitin that mediates the surface turnover of MHC class II (MHCII) and CD86 and plays an essential role in restraining an exhaustion-like program of effector CD4+ T cells [79]. MARCHF1 also regulates type I interferon signaling, T cell activation, and IFN-γ production during infections [80]. Moreover, deletion of the chromosomal positions chr4:164,703,186-165,032,803 encompassing MARCH1 resulted in a previously unreported growth failure, developmental and speech delays and aggressive behavior [81]. It also causes an inflamed tumor microenvironment and suggested as an immune status biomarker for effectiveness of immunotherapy in cancer treatment [82]. We were able to predict the isoforms of *MARCHF1* that are expressed in MS lesions and NAWM; however, utilization of long read sequencing methods are necessary to identify the exact transcripts in each tissue. Although myelin sheath breakdown is the hallmark of MS, the exact targeting and degradation mechanism is unknown. Our results may enable the discovery of the myelin degradation process, as alternative splicing has been shown to affect the selective ubiquitination of different protein isoforms [83]. Identification of the isoforms is important to understand the mechanism, prevent MS progression, and reverse neuronal damage. However, as noted, further studies require more specific methods for the identification of protein isotypes such as IsoSeq and deep proteome sequencing [84, 85].

Both B cell and T cell involvement is known for MS [86]. Our analysis, expectedly, showed the involvement of B-cell mediated immunity in MS lesions. We found brain-derived neurotrophic factor (*BDNF*) as one of the differentially spliced genes that is secreted by B-cells, which prevents axonal loss [87] and highly expressed in the actively demyelinating area [88]. BDNF isoforms have been identified to affect neurogenesis and expression of serotonergic agents [89]. Recent studies show IgG constant region polymorphisms effect antibody stability and dynamics [90]. In MS, a preferential pairing of the IGHV4 gene family with the IGKV1 gene family is shown and IGHV4-39 gene is identified as the most abundant subisotype [91]. Our data showed overexpression of different Ig isotypes and subisotypes in AL and CA lesions. It is important to acknowledge we were able to compare these lesions to NAWM within samples from only 2 donors for CA and 1 donor for AL lesions. However, our results suggest a novel approach in comparison of different categories of lesions could identify specific Ig gene usage and specific biomarkers for molecular classifications via MRI [92].

## CONCLUSION

Our analysis explored various methods and approaches to identify novel molecular characteristics of MS lesions in the brain. We discovered the metabolic pathways were highly affected by gene expression changes. We provided additional insight to differential expression with our differential splicing analysis and identified genes with crucial roles in MS related pathways have alternative isotypes in MS lesions. Our findings identified RNA binding motifs that are involved in these alternative splicing events. Additionally, with the comparison of lesions within donors, we found spontaneous SNPs and alternative isoforms in genes that are essential factors in autoimmunity, neuron homeostasis, and myelination that may have pivotal roles in MS lesions. Overall, our results indicate splice variants in specific MS lesions may be used as biomarkers determine the staging of the lesions as well as treatment targets.

## Data Availability

All data produced in the present study are available upon reasonable request to the authors

https://www.ncbi.nlm.nih.gov/geo/query/acc.cgi?acc=GSE138614

## ACKNOWLEDGEMENTS

We wish to thank members of the KY INBRE Bioinformatics Core and the SWRM lab members for their helpful insight and feedback.

## DATA AVAILABILITY

The samples utilized in this analysis are publicly available in GEO accession GSE138614.

## AUTHOR CONTRIBUTIONS

MS: conceptualization, formal analysis, investigation, methodology, software, validation, visualization, writing – original draft, and writing – review and editing. JHC: formal analysis, software, supervision, and writing – review & editing. JWP: formal analysis and writing – review & editing. ECR: conceptualization, formal analysis, funding acquisition, investigation, methodology, project administration, resources, supervision, and writing – review & editing.

