## Supplemental Tables and Figures for "Gene expression and alternative splicing analysis in a large-scale Multiple Sclerosis study"

**Supplemental Table 1: Top 50 overexpressed genes in sample S6 AL vs NAWM**

| Top 50 overexpressed genes in S6 AL vs NAWM | | | | | |
|---|---|---|---|---|---|
| **ENSEMBL** | **baseMean** | **log2FoldChange** | **pvalue** | **padj** | **Gene ID** |
| ENSG00000211598 | 61.40887189 | 3.358073346 | 9.15498E-11 | 1.67866E-06 | *IGKV4-1* |
| ENSG00000206199 | 17.75290812 | 2.943291209 | 1.46939E-09 | 1.34714E-05 | *ANKUB1* |
| ENSG00000039068 | 120.0125257 | -1.797361702 | 4.15158E-08 | 0.000212318 | *CDH1* |
| ENSG00000145703 | 254.8982805 | 1.962149708 | 4.64985E-08 | 0.000212318 | *IQGAP2* |
| ENSG00000177575 | 1004.400271 | 1.898886006 | 5.78965E-08 | 0.000212318 | *CD163* |
| ENSG00000105877 | 195.3092102 | 2.47680826 | 3.62111E-07 | 0.001106612 | *DNAH11* |
| ENSG00000211934 | 38.80408951 | 2.578669624 | 4.76593E-07 | 0.001248402 | *IGHV1-2* |
| ENSG00000118492 | 43.29878969 | 2.574905533 | 6.53957E-07 | 0.001498869 | *ADGB* |
| ENSG00000162643 | 71.19196462 | 2.355560306 | 7.77243E-07 | 0.001583503 | *DNAI3* |
| ENSG00000034239 | 48.54954016 | 2.398740277 | 1.40438E-06 | 0.002575066 | *CLXN* |
| ENSG00000272398 | 379.1515392 | 2.367795278 | 1.74291E-06 | 0.00290527 | *CD24* |
| ENSG00000133665 | 13.49126362 | 2.4403432 | 2.48329E-06 | 0.003794464 | *DYDC2* |
| ENSG00000224373 | 59.90081173 | 2.239075798 | 3.029E-06 | 0.004272287 | *IGHV4-59* |
| ENSG00000120262 | 125.5207639 | 1.468712179 | 4.4085E-06 | 0.005773881 | *CCDC170* |
| ENSG00000260314 | 95.63672564 | 1.865432839 | 6.34938E-06 | 0.007761484 | *MRC1* |
| ENSG00000065328 | 7.126236149 | -2.298081247 | 8.58088E-06 | 0.007866949 | *MCM10* |
| ENSG00000091181 | 15.62490037 | 2.307878752 | 8.28466E-06 | 0.007866949 | *IL5RA* |
| ENSG00000121966 | 222.2816979 | 2.038563615 | 7.31338E-06 | 0.007866949 | *CXCR4* |
| ENSG00000155761 | 139.5869347 | 2.249632431 | 7.35414E-06 | 0.007866949 | *SPAG17* |
| ENSG00000265401 | 20.95017124 | -2.315901992 | 7.83144E-06 | 0.007866949 | |
| ENSG00000160401 | 39.1306099 | 2.213757402 | 1.65394E-05 | 0.014441233 | *CFAP157* |
| ENSG00000164694 | 20.28898812 | 2.214878451 | 1.88391E-05 | 0.015701569 | *FNDC1* |
| ENSG00000145423 | 153.4475006 | 1.928332026 | 2.3787E-05 | 0.018173263 | *SFRP2* |
| ENSG00000197748 | 368.1429539 | 1.758839983 | 2.29827E-05 | 0.018173263 | *CFAP43* |
| ENSG00000102174 | 77.1954467 | 1.666628837 | 3.64332E-05 | 0.019627304 | *PHEX* |
| ENSG00000106483 | 81.36502251 | 1.820031104 | 3.78071E-05 | 0.019627304 | *SFRP4* |
| ENSG00000111834 | 56.11493438 | 1.821465689 | 3.52117E-05 | 0.019627304 | *RSPH4A* |
| ENSG00000112539 | 49.56403161 | 2.03722544 | 3.76505E-05 | 0.019627304 | *C6orf118* |
| ENSG00000121207 | 209.6606966 | 1.734271623 | 3.58226E-05 | 0.019627304 | *LRAT* |
| ENSG00000165309 | 132.3750071 | 1.739873545 | 3.47326E-05 | 0.019627304 | *ARMC3* |
| ENSG00000165923 | 35.14925521 | 1.937189687 | 3.50647E-05 | 0.019627304 | *AGBL2* |
| ENSG00000168658 | 73.5565266 | 1.915236614 | 3.85353E-05 | 0.019627304 | *VWA3B* |
| ENSG00000168685 | 23.73384978 | 1.90445662 | 2.89403E-05 | 0.019627304 | *IL7R* |
| ENSG00000188452 | 18.46208388 | 2.137244563 | 3.26284E-05 | 0.019627304 | *CERKL* |
| ENSG00000211644 | 68.1204064 | 2.147884521 | 3.31446E-05 | 0.019627304 | *IGLV1-51* |
| ENSG00000213085 | 72.44294647 | 2.130650678 | 3.18533E-05 | 0.019627304 | *CFAP45* |
| ENSG00000158486 | 29.59926372 | 2.071210978 | 4.58878E-05 | 0.022740485 | *DNAH3* |
| ENSG00000007174 | 332.7289818 | 1.824139823 | 5.08799E-05 | 0.023921384 | *DNAH9* |
| ENSG00000163359 | 66.03983974 | 1.526685037 | 4.99738E-05 | 0.023921384 | *COL6A3* |
| ENSG00000140795 | 58.38848635 | 2.087074728 | 5.55412E-05 | 0.02546008 | *MYLK3* |
| ENSG00000132465 | 98.02114091 | 2.056679823 | 7.09261E-05 | 0.030244212 | *JCHAIN* |
| ENSG00000163071 | 30.10427425 | 1.957539826 | 6.86995E-05 | 0.030244212 | *SPATA18* |
| ENSG00000197826 | 17.95429019 | 2.056593967 | 7.08249E-05 | 0.030244212 | *CFAP299* |
| ENSG00000082175 | 84.72871611 | 1.637327291 | 7.909E-05 | 0.03230815 | *PGR* |
| ENSG00000153071 | 825.636917 | 1.105004773 | 7.92903E-05 | 0.03230815 | *DAB2* |
| ENSG00000077327 | 72.8869733 | 1.876823902 | 8.61824E-05 | 0.034301366 | *SPAG6* |
| ENSG00000138061 | 221.8922056 | 1.460920801 | 8.79234E-05 | 0.034301366 | *CYP1B1* |
| ENSG00000133800 | 126.3202893 | 1.797290483 | 9.24253E-05 | 0.035306462 | *LYVE1* |
| ENSG00000134028 | 19.09794775 | 1.918127559 | 9.69426E-05 | 0.03599465 | *ADAMDEC1* |
| ENSG00000145623 | 456.3828503 | 1.266791274 | 9.81529E-05 | 0.03599465 | *OSMR* |

**Supplemental Table 2: Top 50 overexpressed genes in sample S9 CA vs NAWM**

| **Top 50 overexpressed genes S9 CA vs NAWM** | | | | | |
|---|---|---|---|---|---|
| **ENSEMBL** | **baseMean** | **log2FoldChange** | **pvalue** | **padj** | **Gene ID** |
| ENSG00000245067 | 75.4529339 | 2.49552572 | 1.3459E-08 | 0.000286957 | *IGFBP7-AS1* |
| ENSG00000279690 | 59.3808041 | 2.54216831 | 2.2532E-08 | 0.000286957 | |
| ENSG00000039537 | 29.191478 | 2.53304356 | 4.4555E-07 | 0.003782891 | *C6* |
| ENSG00000106823 | 378.396797 | 1.78333132 | 2.5543E-06 | 0.016265411 | *ECM2* |
| ENSG00000047457 | 3159.35988 | 1.76162406 | 5.3687E-06 | 0.027349159 | *CP* |
| ENSG00000272398 | 379.151539 | 2.1292006 | 8.45E-06 | 0.035871659 | *CD24* |
| ENSG00000019991 | 218.899378 | 1.6211616 | 1.05E-05 | 0.038205544 | *HGF* |
| ENSG00000132465 | 98.0211409 | 2.16128719 | 2.4558E-05 | 0.078189431 | *JCHAIN* |
| ENSG00000101916 | 70.3060999 | 1.60035967 | 3.4587E-05 | 0.088097017 | *TLR8* |
| ENSG00000163885 | 48.0416741 | 1.90746442 | 3.3658E-05 | 0.088097017 | *CFAP100* |
| ENSG00000198774 | 183.473235 | 1.81911382 | 3.9802E-05 | 0.092164378 | *RASSF9* |
| ENSG00000165084 | 404.825742 | 1.46861666 | 6.9788E-05 | 0.125262588 | *C8orf34* |
| ENSG00000171659 | 685.159672 | 1.53473049 | 7.3768E-05 | 0.125262588 | *GPR34* |
| ENSG00000179813 | 26.8512811 | 2.0572326 | 6.508E-05 | 0.125262588 | *FAM216B* |
| ENSG00000197747 | 573.33686 | 1.21219009 | 7.3612E-05 | 0.125262588 | *S100A10* |
| ENSG00000164294 | 40.2708332 | 1.71335449 | 8.3114E-05 | 0.13231182 | *GPX8* |
| ENSG00000155761 | 139.586935 | 1.90722347 | 9.4057E-05 | 0.140924895 | *SPAG17* |
| ENSG00000105877 | 195.30921 | 1.78805 | 0.00013976 | 0.197766197 | *DNAH11* |
| ENSG00000152936 | 94.529752 | 1.46055433 | 0.00014902 | 0.199776156 | *LMNTD1* |
| ENSG00000132554 | 280.249723 | 1.58123877 | 0.00016644 | 0.201870249 | *RGS22* |
| ENSG00000135046 | 667.308119 | 1.33696561 | 0.0001624 | 0.201870249 | *ANXA1* |
| ENSG00000248801 | 53.9431494 | 1.83591562 | 0.00022309 | 0.258292626 | *C8orf34-AS1* |
| ENSG00000116745 | 79.8162758 | 1.55862278 | 0.00023497 | 0.260208558 | *RPE65* |
| ENSG00000226690 | 13.675516 | 1.8932191 | 0.00024523 | 0.260263452 | |
| ENSG00000177575 | 1004.40027 | 1.18332513 | 0.00026127 | 0.266197158 | *CD163* |
| ENSG00000188596 | 583.13856 | 1.46285267 | 0.00027892 | 0.273242354 | *CFAP54* |
| ENSG00000006747 | 1107.0058 | 1.17989141 | 0.00029919 | 0.28224635 | *SCIN* |
| ENSG00000173947 | 178.034076 | 1.3409565 | 0.00032454 | 0.295228543 | *PIFO* |
| ENSG00000160838 | 7.58217851 | 1.85517445 | 0.00033689 | 0.295895033 | *LRRC71* |
| ENSG00000154188 | 411.1145 | 1.26110254 | 0.00040363 | 0.332181734 | *ANGPT1* |
| ENSG00000213085 | 72.4429465 | 1.7800244 | 0.00040429 | 0.332181734 | *CFAP45* |
| ENSG00000162643 | 71.1919646 | 1.62116173 | 0.00044103 | 0.340404863 | *DNAI3* |
| ENSG00000270550 | 16.9263375 | 1.79606708 | 0.00043157 | 0.340404863 | *IGHV3-30* |
| ENSG00000162645 | 662.148052 | 1.22378097 | 0.00048533 | 0.363584066 | *GBP2* |
| ENSG00000026025 | 9091.18189 | 1.18933157 | 0.00052779 | 0.37342937 | *VIM* |
| ENSG00000147145 | 96.5266732 | 1.47668401 | 0.0005159 | 0.37342937 | *LPAR4* |
| ENSG00000169385 | 47.4653304 | 1.39948759 | 0.00055438 | 0.381635953 | *RNASE2* |
| ENSG00000140030 | 148.451651 | 1.40950832 | 0.00057138 | 0.382988571 | *GPR65* |
| ENSG00000121316 | 89.4938444 | 1.37074309 | 0.00059566 | 0.389029666 | *PLBD1* |
| ENSG00000175077 | 33.4541282 | 1.48679856 | 0.00062766 | 0.399679032 | *RTP1* |
| ENSG00000205835 | 80.1600578 | 1.61827179 | 0.00066246 | 0.411550536 | *GMNC* |
| ENSG00000118492 | 43.2987897 | 1.73196732 | 0.00073199 | 0.43724422 | *ADGB* |
| ENSG00000144354 | 29.7283792 | 1.4843744 | 0.00073815 | 0.43724422 | *CDCA7* |
| ENSG00000092969 | 911.26171 | 1.09600535 | 0.00103678 | 0.586838719 | *TGFB2* |
| ENSG00000132965 | 335.138721 | 1.22315201 | 0.00103042 | 0.586838719 | *ALOX5AP* |
| ENSG00000110077 | 605.814433 | 0.93687913 | 0.00111428 | 0.616996806 | *MS4A6A* |
| ENSG00000136918 | 11.637692 | 1.67589829 | 0.0011729 | 0.635638922 | *WDR38* |
| ENSG00000034239 | 48.5495402 | 1.53174235 | 0.0016218 | 0.649329941 | *CLXN* |
| ENSG00000091181 | 15.6249004 | 1.65460838 | 0.0013926 | 0.649329941 | *IL5RA* |
| ENSG00000141232 | 1300.53716 | 1.0242855 | 0.00159234 | 0.649329941 | *TOB1* |

**Supplemental Table 3: Top 50 overexpressed in sample S14 CA vs NAWM**

| **Top 50 overexpressed in S14 CA vs NAWM** | | | | | |
|---|---|---|---|---|---|
| **ENSEMBL** | **baseMean** | **log2FoldChange** | **pvalue** | **padj** | **Gene ID** |
| ENSG00000211592 | 1784.875869 | 3.998796543 | 6.64876E-22 | 1.18188E-17 | *IGKC* |
| ENSG00000211938 | 173.3356138 | 3.89399377 | 5.25274E-20 | 4.66864E-16 | *IGHV3-7* |
| ENSG00000163885 | 89.6152127 | 3.167514495 | 2.14961E-17 | 1.27371E-13 | *CFAP100* |
| ENSG00000272398 | 580.5446218 | 3.377801002 | 5.92424E-17 | 2.63273E-13 | *CD24* |
| ENSG00000124491 | 881.320999 | 2.515192517 | 7.92173E-16 | 2.81633E-12 | *F13A1* |
| ENSG00000205835 | 110.0274908 | 3.032773349 | 1.03881E-15 | 3.07766E-12 | *GMNC* |
| ENSG00000132465 | 183.2699715 | 3.374014288 | 3.00556E-15 | 6.67835E-12 | *JCHAIN* |
| ENSG00000165457 | 90.00210278 | 2.680035629 | 2.86533E-15 | 6.67835E-12 | *FOLR2* |
| ENSG00000211896 | 3116.741877 | 3.285655274 | 3.52127E-15 | 6.95491E-12 | *IGHG1* |
| ENSG00000153347 | 73.40745085 | 3.297121024 | 1.08676E-14 | 1.93182E-11 | *FAM81B* |
| ENSG00000198774 | 221.0552752 | 2.625866715 | 1.3368E-13 | 2.16027E-10 | *RASSF9* |
| ENSG00000157330 | 39.56511708 | 3.187624424 | 1.62883E-13 | 2.41284E-10 | *CFAP107* |
| ENSG00000162598 | 79.23552444 | 3.070921959 | 2.10635E-13 | 2.88018E-10 | *C1orf87* |
| ENSG00000155659 | 435.3265925 | 1.96805898 | 2.34342E-13 | 2.97547E-10 | *VSIG4* |
| ENSG00000105877 | 313.0952069 | 2.897467556 | 3.13872E-13 | 3.59889E-10 | *DNAH11* |
| ENSG00000176601 | 88.19437585 | 3.273236839 | 3.23933E-13 | 3.59889E-10 | *MAP3K19* |
| ENSG00000162643 | 108.5616334 | 2.918555149 | 5.87054E-13 | 6.13851E-10 | *DNAI3* |
| ENSG00000110077 | 885.0564178 | 1.572085188 | 7.37255E-13 | 7.2808E-10 | *MS4A6A* |
| ENSG00000039139 | 276.3754475 | 2.536402591 | 9.06774E-13 | 8.0594E-10 | *DNAH5* |
| ENSG00000110079 | 73.03926301 | 2.266522134 | 8.67094E-13 | 8.0594E-10 | *MS4A4A* |
| ENSG00000131951 | 203.4590144 | 2.627258226 | 1.40901E-12 | 1.19269E-09 | *LRRC9* |
| ENSG00000180638 | 79.11539049 | 2.750475892 | 2.00212E-12 | 1.61772E-09 | *SLC47A2* |
| ENSG00000211959 | 33.5856733 | 3.17511463 | 4.45271E-12 | 3.44136E-09 | *IGHV4-39* |
| ENSG00000120708 | 268.3937247 | 1.959618543 | 4.72194E-12 | 3.49738E-09 | *TGFBI* |
| ENSG00000179813 | 47.94559569 | 3.16872989 | 9.36968E-12 | 6.66222E-09 | *FAM216B* |
| ENSG00000034239 | 109.5636494 | 2.592674302 | 1.68292E-11 | 1.1506E-08 | *CLXN* |
| ENSG00000019991 | 307.154295 | 2.116615888 | 1.92879E-11 | 1.26986E-08 | *HGF* |
| ENSG00000155761 | 212.6678428 | 2.915539573 | 2.72353E-11 | 1.72906E-08 | *SPAG17* |
| ENSG00000177575 | 1216.597264 | 2.05085923 | 4.00058E-11 | 2.45221E-08 | *CD163* |
| ENSG00000179178 | 138.4184481 | -2.195741031 | 5.51482E-11 | 3.26771E-08 | *TMEM125* |
| ENSG00000213085 | 87.44363184 | 2.794111802 | 5.96388E-11 | 3.4198E-08 | *CFAP45* |
| ENSG00000092969 | 1219.196722 | 1.600597204 | 1.05876E-10 | 5.88142E-08 | *TGFB2* |
| ENSG00000133800 | 362.6537023 | 2.024821442 | 1.20388E-10 | 6.48489E-08 | *LYVE1* |
| ENSG00000135046 | 653.9823156 | 2.047012651 | 1.38404E-10 | 7.23607E-08 | *ANXA1* |
| ENSG00000162493 | 550.8112957 | 1.577154422 | 1.7193E-10 | 8.73207E-08 | *PDPN* |
| ENSG00000165168 | 1050.919839 | 1.774258895 | 3.42462E-10 | 1.691E-07 | *CYBB* |
| ENSG00000102383 | 208.5823367 | 1.611675358 | 4.12794E-10 | 1.98319E-07 | *ZDHHC15* |
| ENSG00000118492 | 63.67055029 | 2.834140465 | 7.88765E-10 | 3.67516E-07 | *ADGB* |
| ENSG00000211897 | 31.34432628 | 2.655627485 | 8.06318E-10 | 3.67516E-07 | *IGHG3* |
| ENSG00000144354 | 48.0740965 | 2.172868389 | 1.0314E-09 | 4.47176E-07 | *CDCA7* |
| ENSG00000182329 | 120.8804553 | 2.359375984 | 1.01781E-09 | 4.47176E-07 | *KIAA2012* |
| ENSG00000188931 | 65.82734115 | 2.686985942 | 1.19186E-09 | 5.04439E-07 | *CFAP126* |
| ENSG00000260314 | 142.6738709 | 1.889814309 | 1.97715E-09 | 8.17346E-07 | *MRC1* |
| ENSG00000152611 | 22.29570208 | 2.599267693 | 2.6216E-09 | 1.05912E-06 | *CAPSL* |
| ENSG00000147145 | 140.1420192 | 1.878985241 | 2.792E-09 | 1.1029E-06 | *LPAR4* |
| ENSG00000106823 | 383.8262812 | 2.003490716 | 2.99013E-09 | 1.15246E-06 | *ECM2* |
| ENSG00000166596 | 101.1242328 | 2.609133372 | 3.04711E-09 | 1.15246E-06 | *CFAP52* |
| ENSG00000189184 | 476.5560756 | 1.686477887 | 3.32935E-09 | 1.23297E-06 | *PCDH18* |
| ENSG00000197748 | 459.0950423 | 2.074632538 | 3.7792E-09 | 1.371E-06 | *CFAP43* |
| ENSG00000143297 | 16.11791675 | 2.623139754 | 4.26438E-09 | 1.51607E-06 | *FCRL5* |

**Supplemental Table 4: GSEA analysis of differentially expressed genes in AL vs NAWM S6**

| GIS DETAILS | ES | NES | NOM p-val | FDR q-val | FWER p-val | RANK AT MAX |
|---|---|---|---|---|---|---|
| GOBP_CILIUM_MOVEMENT | 137 | 0.72 | 3.04 | 0 | 0 | 0 |
| GOBP_AXONEME_ASSEMBLY | 78 | 0.76 | 2.93 | 0 | 0 | 0 |
| GOBP_CILIUM_OR_FLAGELLUM_DEPENDENT_CELL_MOTILITY | 104 | 0.7 | 2.78 | 0 | 0 | 0 |
| GOBP_MICROTUBULE_BUNDLE_FORMATION | 106 | 0.68 | 2.73 | 0 | 0 | 0 |
| GOBP_AXONEMAL_DYNEIN_COMPLEX_ASSEMBLY | 34 | 0.82 | 2.71 | 0 | 0 | 0 |
| GOBP_SPERM_MOTILITY | 84 | 0.68 | 2.7 | 0 | 0 | 0 |
| GOBP_EXTRACELLULAR_TRANSPORT | 42 | 0.75 | 2.64 | 0 | 0 | 0 |
| GOBP_REGULATION_OF_CILIUM_MOVEMENT | 24 | 0.82 | 2.46 | 0 | 0 | 0 |
| GOBP_MOTILE_CILIUM_ASSEMBLY | 51 | 0.68 | 2.39 | 0 | 0 | 0 |
| GOBP_REGULATION_OF_MICROTUBULE_BASED_MOVEMENT | 38 | 0.68 | 2.32 | 0 | 0 | 0 |
| GOBP_OUTER_DYNEIN_ARM_ASSEMBLY | 20 | 0.81 | 2.31 | 0 | 0 | 0 |
| GOBP_CILIUM_ORGANIZATION | 382 | 0.5 | 2.31 | 0 | 0 | 0 |
| GOBP_REGULATION_OF_CILIUM_BEAT_FREQUENCY | 15 | 0.84 | 2.23 | 0 | 0 | 0.002 |
| GOBP_MICROTUBULE_BASED_MOVEMENT | 354 | 0.47 | 2.21 | 0 | 0 | 0.003 |
| GOBP_POSITIVE_REGULATION_OF_OSTEOBLAST_DIFFERENTIATION | 64 | 0.59 | 2.19 | 0 | 0.001 | 0.009 |
| GOBP_INNER_DYNEIN_ARM_ASSEMBLY | 15 | 0.85 | 2.19 | 0 | 0.001 | 0.009 |
| GOBP_ZYMOGEN_ACTIVATION | 52 | 0.57 | 2.08 | 0 | 0.003 | 0.047 |
| GOBP_SPERM_FLAGELLUM_ASSEMBLY | 30 | 0.66 | 2.07 | 0 | 0.004 | 0.059 |
| GOBP_CELLULAR_RESPONSE_TO_INTERLEUKIN_1 | 72 | 0.55 | 2.06 | 0 | 0.005 | 0.084 |
| S6_ALvsNAWM_GOBP_neg | | | | | | |
| GOBP_REGULATION_OF_POSTSYNAPTIC_MEMBRANE_POTENTIAL | 122 | -0.5 | -2.45 | 0 | 0.001 | 0.001 |
| GOBP_POSTSYNAPSE_ASSEMBLY | 40 | -0.7 | -2.42 | 0 | 0 | 0.001 |
| GOBP_POSITIVE_REGULATION_OF_EXCITATORY_POSTSYNAPTIC_POTENTIAL | 31 | -0.7 | -2.36 | 0 | 0 | 0.001 |
| GOBP_POSTSYNAPTIC_SPECIALIZATION_ASSEMBLY | 28 | -0.7 | -2.35 | 0 | 0 | 0.001 |
| GOBP_MODULATION_OF_EXCITATORY_POSTSYNAPTIC_POTENTIAL | 44 | -0.6 | -2.33 | 0 | 0 | 0.001 |
| GOBP_CHEMICAL_SYNAPTIC_TRANSMISSION_POSTSYNAPTIC | 106 | -0.5 | -2.3 | 0 | 0 | 0.003 |
| GOBP_POSTSYNAPTIC_SPECIALIZATION_ORGANIZATION | 43 | -0.6 | -2.26 | 0 | 0.001 | 0.012 |
| GOBP_GLUTAMATE_RECEPTOR_SIGNALING_PATHWAY | 48 | -0.6 | -2.24 | 0 | 0.001 | 0.014 |
| GOBP_LIGAND_GATED_ION_CHANNEL_SIGNALING_PATHWAY | 29 | -0.7 | -2.24 | 0 | 0.001 | 0.015 |
| GOBP_VESICLE_MEDIATED_TRANSPORT_IN_SYNAPSE | 209 | -0.5 | -2.23 | 0 | 0.001 | 0.015 |
| GOBP_CALCIUM_ION_REGULATED_EXOCYTOSIS | 60 | -0.5 | -2.21 | 0 | 0.002 | 0.022 |
| GOBP_REGULATION_OF_TRANS_SYNAPTIC_SIGNALING | 426 | -0.4 | -2.19 | 0 | 0.002 | 0.024 |
| GOBP_EXCITATORY_SYNAPSE_ASSEMBLY | 33 | -0.6 | -2.19 | 0 | 0.002 | 0.026 |
| GOBP_NEGATIVE_REGULATION_OF_AXON_EXTENSION_INVOLVED_IN_AXON_GUIDANCE | 26 | -0.7 | -2.18 | 0 | 0.002 | 0.027 |
| GOBP_SYNAPTIC_VESICLE_EXOCYTOSIS | 93 | -0.5 | -2.16 | 0 | 0.002 | 0.033 |
| GOBP_CALCIUM_ION_REGULATED_EXOCYTOSIS_OF_NEUROTRANSMITTER | 20 | -0.7 | -2.16 | 0 | 0.002 | 0.038 |
| GOBP_REGULATION_OF_NEUROTRANSMITTER_RECEPTOR_ACTIVITY | 56 | -0.5 | -2.15 | 0 | 0.002 | 0.041 |
| GOBP_PROTEIN_LOCALIZATION_TO_SYNAPSE | 75 | -0.5 | -2.14 | 0 | 0.002 | 0.051 |
| GOBP_POSTSYNAPTIC_DENSITY_ASSEMBLY | 22 | -0.7 | -2.13 | 0 | 0.003 | 0.06 |
| GOBP_SYNAPTIC_VESICLE_RECYCLING | 79 | -0.5 | -2.11 | 0 | 0.003 | 0.074 |

**Supplemental Table 5: GSEA analysis of differentially expressed genes in CA vs NAWM S9**

| GS DETAILS | ES | NES | NOM p-val | FDR q-val | FWER p-val | RANK AT MAX |
|---|---|---|---|---|---|---|
| GOBP_B_CELL_MEDIATED_IMMUNITY | 156 | 0.65 | 2.99 | 0 | 0 | 0 |
| GOBP_AXONEME_ASSEMBLY | 91 | 0.69 | 2.94 | 0 | 0 | 0 |
| GOBP_ADAPTIVE_IMMUNE_RESPONSE | 498 | 0.55 | 2.91 | 0 | 0 | 0 |
| GOBP_LYMPHOCYTE_MEDIATED_IMMUNITY | 295 | 0.56 | 2.88 | 0 | 0 | 0 |
| GOBP_ADAPTIVE_IMMUNE_RESPONSE_BASED_ON_SOMATIC_<br>RECOMBINATION_OF_IMMUNE_RECEPTORS_BUILT_FROM_<br>IMMUNOGLOBULIN_SUPERFAMILY_DOMAINS | 311 | 0.56 | 2.86 | 0 | 0 | 0 |
| GOBP_CILIUM_MOVEMENT | 175 | 0.59 | 2.77 | 0 | 0 | 0 |
| GOBP_HUMORAL_IMMUNE_RESPONSE_MEDIATED_BY<br>CIRCULATING_IMMUNOGLOBULIN | 45 | 0.73 | 2.69 | 0 | 0 | 0 |
| GOBP_COMPLEMENT_ACTIVATION | 52 | 0.7 | 2.66 | 0 | 0 | 0 |
| GOBP_LEUKOCYTE_MEDIATED_IMMUNITY | 384 | 0.52 | 2.66 | 0 | 0 | 0 |
| GOBP_AXONEMAL_DYNEIN_COMPLEX_ASSEMBLY | 38 | 0.73 | 2.57 | 0 | 0 | 0 |
| GOBP_COMPLEMENT_ACTIVATION_CLASSICAL_PATHWAY | 33 | 0.75 | 2.57 | 0 | 0 | 0 |
| GOBP_MICROTUBULE_BUNDLE_FORMATION | 120 | 0.58 | 2.56 | 0 | 0 | 0 |
| GOBP_EXTRACELLULAR_TRANSPORT | 46 | 0.69 | 2.56 | 0 | 0 | 0 |
| GOBP_PEPTIDE_ANTIGEN_ASSEMBLY_WITH<br>_MHC_PROTEIN_COMPLEX | 18 | 0.86 | 2.56 | 0 | 0 | 0 |
| GOBP_MONOCYTE_CHEMOTAXIS | 50 | 0.66 | 2.54 | 0 | 0 | 0 |
| GOBP_CILIUM_OR_FLAGELLUM_DEPENDENT_CELL_MOTILITY | 136 | 0.56 | 2.53 | 0 | 0 | 0 |
| GOBP_HUMORAL_IMMUNE_RESPONSE<br>GOBP_CELL_KILLING | 160 | 0.53 | 2.49 | 0 | 0 | 0 |
| GOBP_POSITIVE_REGULATION_OF_CHEMOKINE_PRODUCTION | 62 | 0.62 | 2.46 | 0 | 0 | 0 |
| GOBP_ANTIGEN_PROCESSING_AND_PRESENTATION<br>OF_PEPTIDE_ANTIGEN | 68 | 0.61 | 2.46 | 0 | 0 | 0 |
| S9_CAvsNAWM_GOBP_neg | | | | | | |
| GOBP_SHORT_CHAIN_FATTY_ACID_METABOLIC_PROCESS | 15 | -0.72 | -2.02 | 0 | 0.134 | 0.161 |
| GOBP_FATTY_ACID_BETA_OXIDATION | 74 | -0.48 | -1.94 | 0 | 0.201 | 0.391 |
| GOBP_OXIDATIVE_PHOSPHORYLATION | 130 | -0.43 | -1.92 | 0 | 0.178 | 0.483 |
| GOBP_MITOCHONDRIAL_ELECTRON_TRANSPORT_<br>CYTOCHROME_C_TO_OXYGEN | 20 | -0.64 | -1.92 | 0 | 0.137 | 0.492 |
| GOBP_FATTY_ACID_CATABOLIC_PROCESS | 102 | -0.45 | -1.9 | 0 | 0.152 | 0.61 |
| GOBP_ATP_SYNTHESIS_COUPLED_ELECTRON_TRANSPORT | 88 | -0.46 | -1.89 | 0 | 0.153 | 0.682 |
| GOBP_AEROBIC_RESPIRATION | 177 | -0.41 | -1.88 | 0 | 0.151 | 0.735 |
| GOBP_MONOCARBOXYLIC_ACID_CATABOLIC_PROCESS | 123 | -0.42 | -1.86 | 0 | 0.152 | 0.79 |
| GOBP_2_OXOGLUTARATE_METABOLIC_PROCESS | 15 | -0.66 | -1.85 | 0 | 0.158 | 0.842 |
| GOBP_CHEMICAL_SYNAPTIC_TRANSMISSION_POSTSYNAPTIC | 104 | -0.43 | -1.85 | 0 | 0.145 | 0.85 |
| GOBP_INHIBITORY_POSTSYNAPTIC_POTENTIAL | 17 | -0.63 | -1.85 | 0.008 | 0.136 | 0.855 |
| GOBP_REGULATION_OF_PROTON_TRANSPORT | 16 | -0.66 | -1.84 | 0.004 | 0.135 | 0.874 |
| GOBP_REGULATION_OF_TRANSCRIPTION_OF_<br>NUCLEOLAR_LARGE_RRNA_BY_RNA_POLYMERASE_I | 15 | -0.66 | -1.82 | 0 | 0.152 | 0.922 |

**Supplemental Table 6: GSEA analysis of differentially expressed genes in CA vs NAWM S14**

| GS DETAILS | ES | NES | NOM p-val | FDR q-val | FWER p-val | RANK AT MAX |
|---|---|---|---|---|---|---|
| GOBP_AXONEME_ASSEMBLY | 85 | 0.79 | 2.88 | 0 | 0 | 0 |
| GOBP_CILIUM_MOVEMENT | 149 | 0.68 | 2.74 | 0 | 0 | 0 |
| GOBP_MICROTUBULE_BUNDLE_FORMATION | 112 | 0.7 | 2.73 | 0 | 0 | 0 |
| GOBP_AXONEMAL_DYNEIN_COMPLEX_ASSEMBLY | 37 | 0.83 | 2.65 | 0 | 0 | 0 |
| GOBP_B_CELL_MEDIATED_IMMUNITY | 113 | 0.68 | 2.63 | 0 | 0 | 0 |
| GOBP_EXTRACELLULAR_TRANSPORT | 45 | 0.77 | 2.54 | 0 | 0 | 0 |
| GOBP_ADAPTIVE_IMMUNE_RESPONSE_BASED_ON_SOMATIC_ RECOMBINATION_OF_IMMUNE_RECEPTORS_BUILT_FROM_ IMMUNOGLOBULIN_SUPERFAMILY_DOMAINS | 245 | 0.6 | 2.54 | 0 | 0 | 0 |
| GOBP_CILIUM_OR_FLAGELLUM_DEPENDENT_CELL_MOTILITY | 111 | 0.65 | 2.53 | 0 | 0 | 0 |
| GOBP_HUMORAL_IMMUNE_RESPONSE_MEDIATED_BY_CIRCULATING_IMMUNOGLOBULIN | 38 | 0.78 | 2.52 | 0 | 0 | 0 |
| GOBP_MOTILE_CILIUM_ASSEMBLY | 53 | 0.71 | 2.47 | 0 | 0 | 0 |
| GOBP_ADAPTIVE_IMMUNE_RESPONSE | 349 | 0.56 | 2.45 | 0 | 0 | 0 |
| GOBP_LYMPHOCYTE_MEDIATED_IMMUNITY | 220 | 0.58 | 2.43 | 0 | 0 | 0 |
| GOBP_COMPLEMENT_ACTIVATION | 42 | 0.74 | 2.42 | 0 | 0 | 0 |
| GOBP_REGULATION_OF_CILIUM_MOVEMENT | 25 | 0.81 | 2.38 | 0 | 0 | 0 |
| GOBP_COMPLEMENT_ACTIVATION_CLASSICAL_PATHWAY | 30 | 0.78 | 2.38 | 0 | 0 | 0 |
| GOBP_LEUKOCYTE_MEDIATED_IMMUNITY | 294 | 0.54 | 2.33 | 0 | 0 | 0 |
| GOBP_CILIUM_ORGANIZATION | 393 | 0.53 | 2.32 | 0 | 0 | 0 |
| GOBP_HUMORAL_IMMUNE_RESPONSE | 114 | 0.6 | 2.3 | 0 | 0 | 0 |
| GOBP_SPERM_MOTILITY | 86 | 0.63 | 2.29 | 0 | 0 | 0 |
| GOBP_POSITIVE_REGULATION_OF_OSTEOBLAST_DIFFERENTIATION | 59 | 0.66 | 2.29 | 0 | 0 | 0 |
| S14_CAvsNAWM_GOBP_neg | | | | | | |
| GOBP_ACETYL_COA_METABOLIC_PROCESS | 29 | -0.6 | -2.07 | 0 | 0.101 | 0.117 |
| GOBP_OLIGODENDROCYTE_DEVELOPMENT | 46 | -0.5 | -2.05 | 0 | 0.065 | 0.146 |
| GOBP_GPI_ANCHOR_METABOLIC_PROCESS | 30 | -0.6 | -2.04 | 0 | 0.047 | 0.158 |
| GOBP_ENSHEATHMENT_OF_NEURONS | 135 | -0.4 | -1.98 | 0 | 0.079 | 0.315 |
| GOBP_AXON_ENSHEATHMENT_IN_CENTRAL_NERVOUS_SYSTEM | 25 | -0.6 | -1.95 | 0 | 0.102 | 0.468 |
| GOBP_STEROL_METABOLIC_PROCESS | 119 | -0.4 | -1.93 | 0 | 0.106 | 0.543 |
| GOBP_STEROL_BIOSYNTHETIC_PROCESS | 56 | -0.5 | -1.93 | 0 | 0.091 | 0.544 |
| GOBP_OLIGODENDROCYTE_DIFFERENTIATION | 97 | -0.4 | -1.88 | 0 | 0.132 | 0.723 |
| GOBP_RESPONSE_TO_IMMOBILIZATION_STRESS | 16 | -0.6 | -1.8 | 0.011 | 0.259 | 0.942 |
| GOBP_THIOESTER_METABOLIC_PROCESS | 77 | -0.4 | -1.79 | 0 | 0.245 | 0.951 |
| GOBP_REGULATION_OF_CAMP_MEDIATED_SIGNALING | 20 | -0.6 | -1.74 | 0 | 0.362 | 0.992 |
| GOBP_TAIL_ANCHORED_MEMBRANE_PROTEIN_INSERTION_INTO_ER_MEMBRANE | 17 | -0.6 | -1.72 | 0.005 | 0.392 | 0.997 |
| GOBP_N_ACETYLGLUCOSAMINE_METABOLIC_PROCESS | 17 | -0.6 | -1.71 | 0.018 | 0.419 | 0.999 |
| GOBP_RRNA_METHYLATION | 27 | -0.5 | -1.68 | 0.011 | 0.498 | 1 |
| GOBP_THIOESTER_BIOSYNTHETIC_PROCESS | 41 | -0.4 | -1.68 | 0.006 | 0.474 | 1 |
| GOBP_SENSORY_PERCEPTION_OF_BITTER_TASTE | 21 | -0.5 | -1.67 | 0.006 | 0.475 | 1 |
| GOBP_DETECTION_OF_CHEMICAL_STIMULUS_INVOLVED _IN_SENSORY_PERCEPTION_OF_TASTE | 20 | -0.5 | -1.66 | 0.013 | 0.463 | 1 |
| GOBP_TRNA_PROCESSING | 133 | -0.4 | -1.66 | 0 | 0.444 | 1 |
| GOBP_NUCLEOSIDE_BISPHOSPHATE_BIOSYNTHETIC_PROCESS | 52 | -0.4 | -1.66 | 0.003 | 0.431 | 1 |
| GOBP_NEGATIVE_REGULATION_OF_TOR_SIGNALING | 68 | -0.4 | -1.64 | 0.007 | 0.492 | 1 |

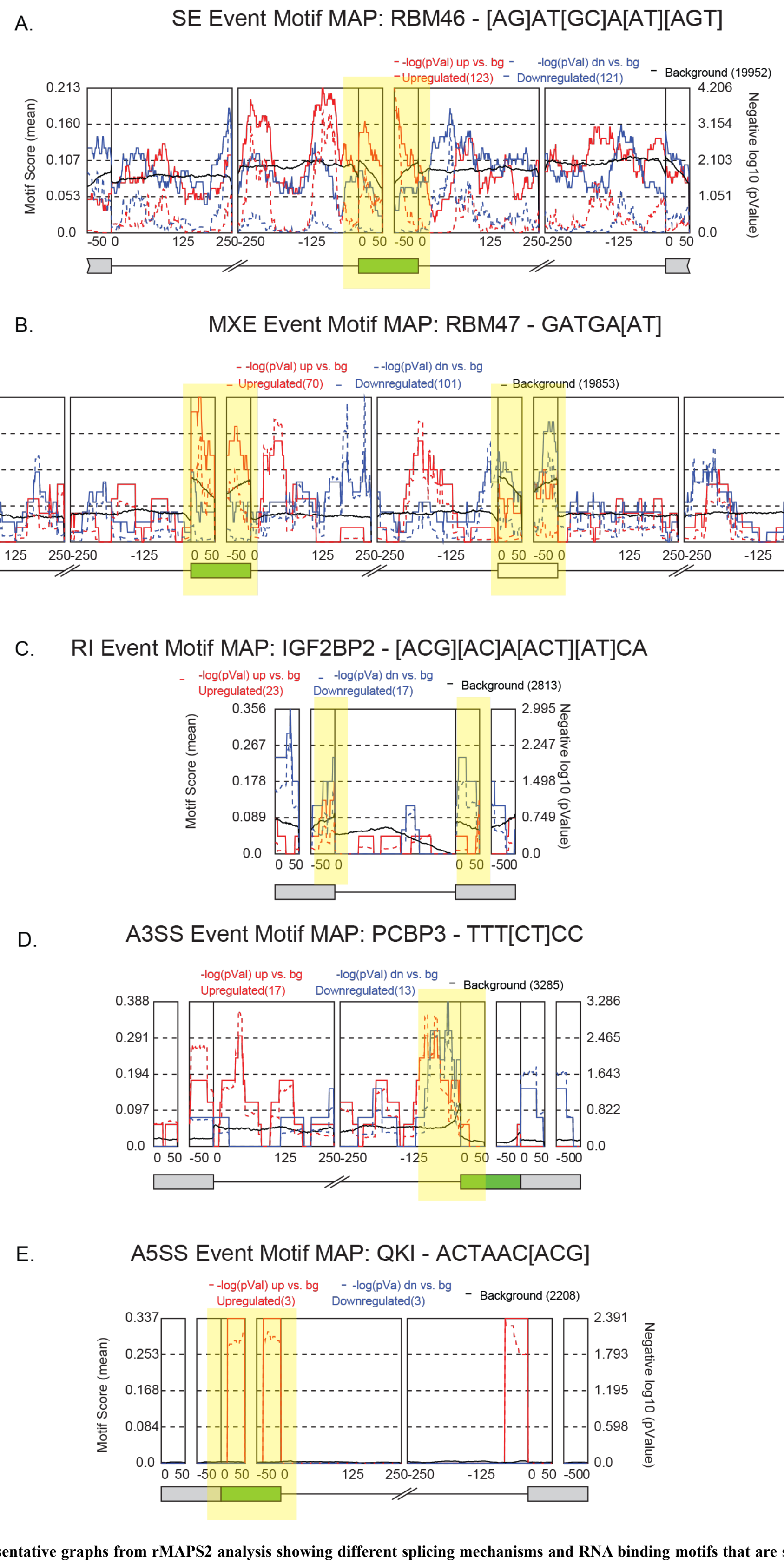


**Supplemental Figure 1: Representative graphs from rMAPS2 analysis showing different splicing mechanisms and RNA binding motifs that are significantly involved in differential splicing of MS and non-MS tissues A.** Skipped Exon (SE), **B.** Mutually Exclusive Exons (MXE), **C.** Retained Intron (RI), **D.** Alternative 3' Splice Site (A3SS), **E.** Alternative 5' Splice Site (A5SS) events. Enriched RBP motifs are highlighted.

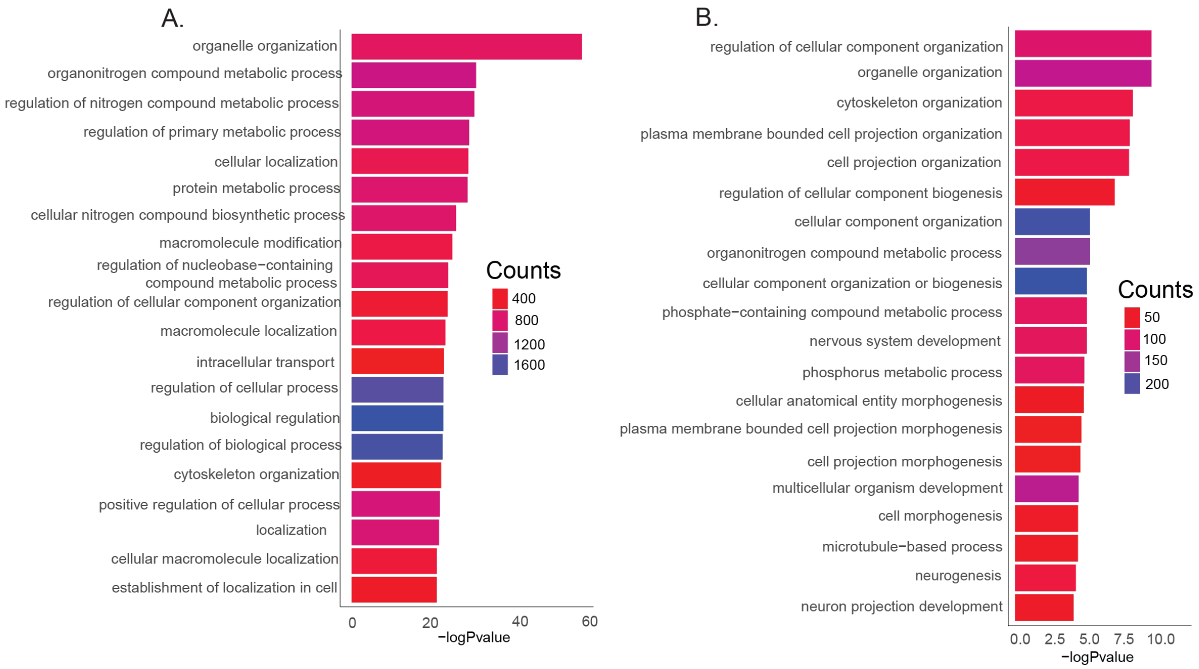


**Supplemental Figure 2: Gene ontology biological processes enrichment graphs of differentially spliced genes in A.** AL compared to NAWM from donor S6 and **B.** CA compared to NAWM from donors S9 and S14.
